# Mapping the Antimicrobial Susceptibility of Methicillin-Resistant *Staphylococcus aureus* in Western Ethiopia: A multicenter cross-sectional study

**DOI:** 10.64898/2026.03.05.26347706

**Authors:** Endalu Tesfaye Guteta, Abdi Diriba, Kume Tesfaye, Essa Kedir, Milkias Wakgari, Demeke Jabessa, Motuma Chali, Keresa Biyena, Getu Sileshi, Girmaye Jobir

**Affiliations:** Nekemte Public Health Research and Referral Laboratory Center, Nekemte, Ethiopia; Armaeur Hansen Research Institute, Addis Ababa, Ethiopia; Department of Medical Laboratory Sciences, Institute of Health Sciences, Wollega University, Nekemte, Ethiopia

**Keywords:** Methicillin-resistant *S. aureus*, antimicrobial resistance, multidrug resistant MRSA

## Abstract

From 2021 to 2025, MRSA emerged as a major multidrug-resistant pathogen in the study area. Among 545 S. aureus isolates, 67.2% were MRSA, disproportionately affecting children under five (26.5%) and males (55.5%). Case incidence more than doubled by 2025, suggesting rising transmission or resistance. Most isolates were hospital-associated (85.2%), predominantly from outpatients (88.5%), with middle ear discharge as the main source (67%). Gentamicin showed the highest susceptibility (72.1%), while penicillin G resistance was nearly universal (96.7%). The majority (93.4%) were multidrug-resistant, with high MARI values indicating widespread and likely inappropriate antibiotic use. These findings reflect a complex interplay between pathogen behavior, antimicrobial use, and healthcare practices. Increasing MRSA burden may stem from inadequate infection control, poor stewardship, or enhanced community transmission. Incorporating molecular typing could deepen understanding of strain diversity and resistance mechanisms to guide targeted interventions

**Importance:** To address scarce antimicrobial resistance data undermining patient care, this first multicenter study maps MRSA susceptibility in Western Ethiopia. Findings guide empiric therapy, establish a stewardship baseline, and contribute to global surveillance. Widespread multidrug resistance and high resistance indices reflect strong antimicrobial pressure and misuse, underscoring the urgent need for antimicrobial stewardship, surveillance, and infection control.

## Introduction

*Staphylococcus aureus* is a gram-positive, catalase and coagulase-positive commensal bacterium that colonizes the skin and nares of approximately 30% of individuals (1). It inhabits moist mucous membranes and can cause invasive infections if it enters sterile areas (2). Up to 100% of patients experience transient nasal carriage, with many infections arising from prior colonization (3). *S. aureus* causes serious diseases, including meningitis, septicemia, pneumonia, endocarditis, and osteomyelitis (4,5), demonstrating exceptional adaptability through antibiotic resistance (6).

Methicillin-resistant *Staphylococcus aureus* (MRSA) is a strain resistant to β-lactam antibiotics (penicillins and cephalosporins), making infections difficult to treat due to the associated multidrug resistance (7,8). Resistance is mediated by the *mecA* gene, which produces an altered penicillin-binding protein (PBP2a) with a reduced affinity for β-lactams (9–11). Epidemiologically, MRSA is classified as community-associated (CA-MRSA), healthcare-associated (HA-MRSA), or livestock-associated (LA-MRSA) (12–18). CA-MRSA is typically susceptible to non-β-lactam antibiotics, whereas HA-MRSA is often multidrug resistant (19).

MRSA is a global health threat linked to 64% higher mortality than non-resistant infections and is listed as a high-priority pathogen by the WHO (20,21). It is highly prevalent in Europe and the U.S. and is responsible for >50% of hospital-acquired infections (22). MRSA affects approximately 29% of carriers, causing prolonged hospital stays, increased costs, and thousands of deaths globally, with an estimated 171,000 invasive infections annually in Europe alone (23,24). Treatment is limited to a few agents (vancomycin and teicoplanin) because of resistance (25). In sub-Saharan Africa, limited surveillance data show MRSA prevalence ranging from 1.25% to 46% (26), while in East Africa, it is 53.4% (27). In Ethiopia, the prevalence varies widely: 68.5% in Gondar (28), 36.9% in Debre Markos (29), 7.5% in Addis Ababa (30), 11.2% in Harar (31), and 17.9% in Hawassa (32). Due to the limited number of short-term studies, this five-year assessment (2021–2025) examined the prevalence of MRSA and its antimicrobial susceptibility in western Ethiopia

## Methods and materials

### Study area, design and period

This laboratory-based analysis was conducted at the Nekemte Public Health Research and Referral Laboratory Center, a regional facility in the East Wallaga Zone of the Oromia Region. The center provides diagnostic, research, and mentorship services to catchment health facilities and serves as an AMR surveillance site in western Ethiopia. Clinical specimens collected from six hospitals, three health centers, and six medium-sized clinics between January 1, 2021, and December 31, 2025, were included.

### Data collection

All clinical specimens positive for *S. aureus* were collected. Patient demographic characteristics (age and sex), type of referring health facilities (hospitals, health centers, and clinics), patient location (outpatient or inpatient), specimen type, and antimicrobial susceptibility profiles.

### Clinical specimen processing, *S. aureus* isolation and identification

Clinical specimens, including ear discharge, blood, urine, pus, body fluids, sputum, and nasal swabs, from healthcare facilities meeting the acceptance criteria were cultured on blood and mannitol salt agar at 37°C for 24 h. Presumptive *S. aureus*—identified by golden-yellow colonies on mannitol salt agar and beta-hemolysis on blood agar—was confirmed via Gram staining, catalase, and coagulase tests.

### Operational definitions

Methicillin-resistant *S. aureus*: *S. aureus* isolates with a cefoxitin zone diameter ≤21 mm on Mueller–Hinton agar after 16–18 hours of incubation (29).

Multidrug resistance: Non-susceptibility to at least one antimicrobial agent in three or more distinct antimicrobial classes (33).

Multiple antibiotic resistance index: Calculated for each isolate by dividing the number of antibiotics to which the isolate was resistant by the total number of antibiotics tested (34–36).

### Antimicrobial susceptibility testing

Methicillin resistance was determined using the cefoxitin (30 µg) disk diffusion method according to CLSI guidelines, with zone diameters of ≥22 mm interpreted as susceptible and ≤21 mm as MRSA. MRSA isolates were tested on Mueller–Hinton agar using a 0.5 McFarland suspension incubated at 37°C for 24 h. The antibiotics tested included penicillin G (10 IU), cefoxitin (30 µg), clindamycin (2 µg), cotrimoxazole (25 µg), azithromycin (15 µg), gentamicin (10 µg), ciprofloxacin (5 µg), chloramphenicol (30 µg), tetracycline (30 µg), and nitrofurantoin (300 µg).

### Quality assurance

All culture media and antimicrobial disks were routinely quality-checked using CLSI guidelines and ATCC reference strains (*E. coli* 25922, *P. aeruginosa* 27853, *S. aureus* 25923) to ensure accurate and reliable testing.

### Data analysis and interpretation

Data completeness was assessed using the standard reporting feature, and records with missing data were excluded. Antimicrobial susceptibility patterns were analyzed by determining proportions of resistant, intermediate, and susceptible isolates. Sociodemographic and susceptibility data were entered into Epi Data version 4.6 and exported to SPSS for analysis.

## Results

### Sociodemographic characteristics and frequency of MRSA isolation

The patients ranged in age from 1 to 82 years (mean ± SD: 21.5 ± 17.5 years). Of the 2,742 specimens collected, 1,419 (51.8%) yielded bacterial growth. Among these, 545 (38.4%) were *S. aureus* and 366 (67.2%) were MRSA. A substantial proportion of MRSA isolates (26.5%) were from children under five years of age, and the isolates were more frequent in males (55.5%), highlighting potential demographic trends (Table 1).

**Table 1:** Frequency of methicillin-resistant *S. aureus* with respect to sociodemographic variables at Western Ethiopia, 2021-2025.

| Variable | Category | Number of MRSA (n = 366) |  |
| --- | --- | --- | --- |
|  |  | Frequency | Percentage |
| Age | <5 | 97 | 26.5 |
|  | 5-14 | 47 | 12.8 |
|  | 15-24 | 64 | 17.5 |
|  | 25-34 | 75 | 20.5 |
| | $\geq 35$ | 83 | 22.7 |
| Sex | Male | 203 | 55.5 |
|  | Female | 163 | 44.5 |
| Type of referring health facility | Hospital | 312 | 85.2 |
|  | Health center | 21 | 5.7 |
|  | Clinic | 33 | 9.1 |
| Patient location | Outpatient department | 324 | 88.5 |
|  | Inpatient department | 42 | 11.5 |
| Type of clinical specimen | Middle ear discharge | 245 | 67.0 |
|  | Blood | 32 | 8.7 |
|  | Urine | 26 | 7.1 |
|  | Pus | 56 | 15.3 |
|  | Body fluids | 4 | 1.1 |
|  | Sputum | 2 | 0.5 |
|  | Nasal swab | 1 | 0.3 |

### Trend of MRSA over the five years

Annual MRSA cases rose steadily from 2021 to 2025, increasing from 49 to 60 in 2022 (22.4% rise), then to 75 in 2023, followed by a modest rise to 80 in 2024, before surging to a peak of 102 in 2025 (Figure 1).

**Figure 1:**
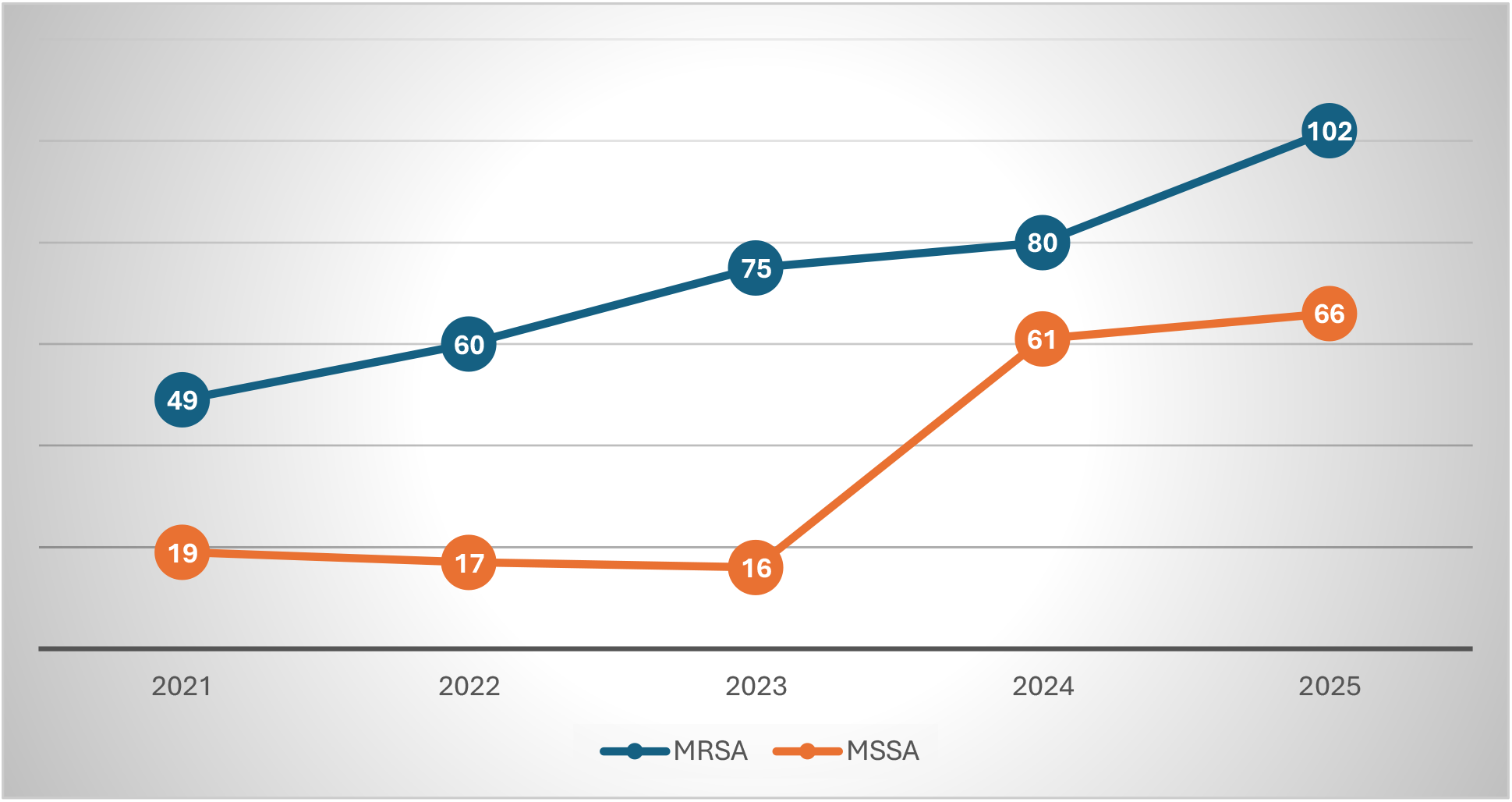
Trends of MRSA over five years at Western Ethiopia, 2021-2025.

### Antimicrobial susceptibility patterns of MRSA

Antimicrobial susceptibility testing of MRSA isolates using the disk diffusion method revealed notable variability across the antibiotics tested. The highest susceptibility was observed for gentamicin 264 (72.1%), followed by chloramphenicol 189 (51.6%) and clindamycin 184 (50.3%). In contrast, the majority of MRSA isolates were resistant to penicillin G 354 (96.7%) and tetracycline 262 (71.6%), highlighting a high level of resistance to commonly used antibiotics (Table 2).

**Table 2:** Antimicrobial susceptibility patterns of MRSA at Western Ethiopia, 2021-2025.

| Antibiotics tested | Susceptibility test results |  |  |
| --- | --- | --- | --- |
|  | Susceptible | Intermediate | Resistant |
| Gentamicin | 264 (72.1) | 13 (3.6) | 89 (24.3) |
| Ciprofloxacin | 152 (41.5) | 21 (5.7) | 193 (52.7) |
| Tetracycline | 89 (24.3) | 15 (4.1) | 262 (71.6) |
| Trimethoprim-sulfamethoxazole | 90 (24.6) | 22 (6.0) | 254 (68.4) |
| Chloramphenicol | 189 (51.6) | 9 (2.5) | 168 (45.9) |
| Azithromycin | 106 (29.0) | 16 (4.4) | 244 (66.6) |
| Clindamycin | 184 (50.3) | 34 (9.3) | 148 (40.4) |
| Penicillin G | 12 (3.3) | 0 (0.0) | 354 (96.7) |
| Cefoxitin | 0 (0.0) | 0 (0.0) | 366 (100) |
| Nitrofurantoin* | 8 (2.2) | 0 (0.0) | 14 (3.8) |

\*Tested only for urine isolates

### Multidrug resistant MRSA and Multiple antibiotic resistance index (MARI)

In this study, 342 (93.4%) MRSA isolates were MDR; 68 (19.9%) were resistant to seven agents, and one (0.3%) was resistant to ten agents. The MARI exceeded 0.2 for all MDR isolates, indicating their origin from high-risk environments with substantial antibiotic exposure (Table 3).

**Table 3:** Multidrug resistance and multiple antibiotic resistance index of MRSA isolates at Western Ethiopia, 2021-2025.

| Number of antibiotics which the isolate resisted | Number of antibiotics tested | Number of isolates (%) | MAR index |
| --- | --- | --- | --- |
| 1 | 9 | 3 (0.9) | 0.1 |
| 2 | 9 | 21 (6.1) | 0.2 |
| 3 | 9 | 44 (12.9) | 0.3 |
| 4 | 9 | 40 (11.7) | 0.4 |
| 5 | 9 | 56 (16.4) | 0.6 |
| 6 | 9 | 51 (14.9) | 0.7 |
| 7 | 9 | 68 (19.9) | 0.8 |
| 8 | 9 | 56 (16.4) | 0.9 |
| 9 | 9 | 26 (7.6) | 1.0 |
| 10 | 10 | 1 (0.3) | 1.0 |

## Discussion

This study examined the MRSA prevalence, susceptibility patterns, and multidrug resistance over five years. The prevalence of MRSA was 67.2% (95% CI: 63.1–71.0), which is higher than that reported in Addis Ababa (27%) (37), Hawassa (17.9%) (32), Harar (11.2%) (31), Nigeria (35.4%) (38), Tanzania (48.5%) (39), Saudi Arabia (52.7%) (40), Pakistan (49%) (41), and Nepal (46%) (5). This finding was consistent with Gondar (68.5%) (28) but lower than Arba Minch (82.3%) (42) and Somalia (81%) (43), India (73.7%) (44), and Pakistan (80%) (45). These variations reflect differences in study design, location, sample size, patient groups, laboratory methods, and antibiotic use, underscoring the global public health impact of this issue.

Age-wise analysis revealed MRSA predominance in children under five (26.5%), contrasting with studies from Arba Minch (18–28 years) (46), Debre Markos (15–30 years) (47), Addis Ababa (25–34 years) (33), and Nigeria (31–45 years) (48). These differences likely reflect variations in the study populations. However, MRSA was more common in males (55.5% vs. 44.5%), consistent with reports from Gondar (28), Addis Ababa (37), Arba Minch (42), Saudi Arabia (40), and Pakistan (41), although some studies found a female predominance (32,49). No single biological factor explains sex differences; the prevalence likely reflects a mix of biological, behavioral, and environmental factors (50,51).

This study found that 88.5% of MRSA isolates came from outpatients, aligning with one Indian study (52) but contrasting with reports from Addis Ababa (37), Hawassa (32), Gondar (28), and Tanzania (39), where MRSA was more common among inpatients than outpatients. The high outpatient prevalence suggests an increasing burden of CA-MRSA, underscoring the need for updated empirical treatment and community-level interventions. Most isolates were from middle ear discharge (67%), followed by pus (15.3%). This differs from other Ethiopian studies, where pus predominated (20.3% (33) and 77.6% (37), and from studies in Nigeria (50%) (48), India (35.7%) (52), and Pakistan (51%) (45), where pus was the main source. Such discrepancies may reflect differences in colonization sites, patient populations, antibiotic use, clinical procedures or detection methods.

Antibiotic susceptibility testing revealed varying resistance patterns. Sensitivity was highest to gentamicin (72.1%), while 96.7% were resistant to penicillin, consistent with studies reporting 95.7% gentamicin sensitivity and 93.3% penicillin resistance (31), and another study where 50% were gentamicin-sensitive and all were penicillin-resistant (37). Ciprofloxacin resistance was 52.7%, which is similar to that reported in one study (55.3%) (53) but different from that reported in another (86.4%) (54). Over half of the isolates were resistant to trimethoprim-sulfamethoxazole (68.4%), azithromycin (66.6%), and clindamycin (50.3%). Other studies have shown variable patterns globally (33,41,42,49). Multidrug resistance (MDR) was 93.4% (95% CI: 90.6-95.6), exceeding rates in Northwestern Ethiopia (57.1%) (29) and Nigeria (57.7%) (34), but consistent with that in Nepal (94.1%) (55). Discrepancies in MDR MRSA distribution stem from geography, setting, period, dominant clones, antibiotic use, and methods used. The MAR index ranged from 0.1 to 1.0; values >0.2 indicate high-risk sources under strong antibiotic pressure, posing public health concerns (35,36), while ≤0.2 suggests low antibiotic use settings (34).

### Limitations

Molecular identification of resistance genes was not performed, so the genetic basis of observed resistance patterns could not be determined. Additionally, patient- or healthcare-related risk factor data were unavailable, preventing analysis of potential resistance predictors. These gaps limit the depth, comprehensiveness, and generalizability of the findings.

## Conclusion

The high prevalence of MRSA and multidrug resistance severely limits treatment options. Elevated MAR index values indicate prolonged antibiotic pressure from indiscriminate use. These findings underscore the urgent need for strengthened infection control, antimicrobial stewardship, and surveillance.

## Data Availability

All data produced in the present study are available upon reasonable request to the authors

## Acknowledgements

The authors would like to thank Nekemte Public Health Research and Referral Laboratory Center for providing permission to conduct this study.

## Author contributions

Conceptualization: ET, AD, KT, DJ; Funding acquisition: ET, DM, KB; Project administration: ET, DJ, KT, GS; Supervision: KB, GS, DJ, GJ, EK; Writing – original draft: ET, AD, KT; Data; curation: ET, DJ, AD, EK, GJ; Investigation: ET, AD, MC, KT, EK; Resources: ET, GS, DJ, KB; Validation: ET, GS, DJ, AD; Writing – review and editing: EK, AD, GS, KT, KB; Formal analysis – ET; Methodology: ET, AD, GJ, KT, DJ; Software: ET, GS, AD, KT; Visualization: AD, ET, DJ

## Data availability

All the data relevant to this manuscript are available upon request from the corresponding author.

## Ethics approval

This study was approved by Nekemte Public Health Research and Referral Laboratory Center Research Committee (Ref. No: NPHRRL/883/04/18). The research involved the use of previously archived laboratory data. There was no direct interaction with patients, and no identifying personal information (such as names or medical record numbers) was collected by the research team. All data were anonymized prior to analysis to ensure patient confidentiality

